# Changes to Fruit and Vegetable Intake and Waste When Households Receive Free Produce

**DOI:** 10.1101/2025.04.23.25326258

**Authors:** Brian E. Roe, Hanim E. Diktas, Danyi Qi, Corby K. Martin, John W. Apolzan

## Abstract

**Objective:** The study evaluated changes in household food intake, the waste of fruits and vegetables (FV), and FV inventories after supplemental produce was provided free of charge and in response to a smart coaching intervention to reduce food waste and replace less nutritious foods with FV.

**Design:** Households measured food intake and waste for ≥3 days before and after intervention. Households were randomized to receive either an intervention to reduce food waste and replace less healthy foods with FV or a control intervention. Both groups received free FV and measured FV inventories before and after intervention.

**Setting:** Participants were from the Baton Rouge, Louisiana region and picked up FV at a central location.

**Participants:** 46 adults and their household members.

**Results:** Treatment participants increased intake of fruits (0.33 servings/day, p=0.09) and vegetables (0.50 servings/day, p=0.01) compared to the control group. All participants reported a decrease in daily total caloric intake (133 kcal/day, p=0.04), an increase in the number (9.5 events/period, p<0.001) and average magnitude (100.5 g/event, p=0.005) of FV waste events, and an increase in fresh FV inventories (4.1 kg/household, p=0.001) after receiving free FVs. Compared to the control group, treatment participants reported less FV waste during eating occasions (22.2 g/day, p=0.09) and an increase in frozen FV inventories (1.8 kg/household, p=0.04).

**Conclusions:** Providing free FVs without additional intervention does not increase FV intake but does lead to more and larger FV waste events. When coupled with targeted information to improve diet quality and reduce waste, free FV provision can lead to increased FV intake with no significant increase in energy intake or plate waste and smaller increases in the number and magnitude of FV waste events, suggesting that pairing intensive intervention efforts with free FV provision is critical to translate program resources into improved nutrition without increasing waste.

## Introduction

Federal guidelines (USDA & USDHHS 2020) recommend a dietary pattern featuring greater consumption of fruits and vegetables (FV) to reduce the risk of cardiovascular disease, type-2 diabetes, some cancers, and obesity, whose prevention would yield numerous health and economic benefits (Biener et al. 2017). Despite extensive public messaging and well-documented evidence of health and economic benefits, only about 10% of Americans eat the recommended amounts of FV (Lee et al. 2022) with the high cost of FV identified as one barrier to increasing FV intake (Pollard et al. 2002, Yeh et al. 2008).

Several studies identify FV subsidies as an effective approach for increasing FV intake (see An 2012, Black et al. 2012, Gittelsohn et al. 2017, Jabari et al. 2024, Bhat et al. 2021, Burgaz et al. 2023, Steer et al. 2023 and Verghese et al. 2019 for reviews of earlier works and Xie et al. (2021) and Lowery et al. (2022) for more recent studies). FV subsidy programs targeting Supplemental Nutrition Assistance Program (SNAP) participants are supported through programs in recent Farm Bills (Gus Schumacher Nutrition Incentive Program, 2018 Farm Bill, and Food Insecurity Nutrition Incentive Program, 2014 Farm Bill), including $41 million during fiscal 2019 and $536 million over 10 years (Miller et al. 2019). These programs and several foundations fund numerous programs across the country such as Double Up Food Bucks or the Summer Electronic Benefits Transfer for Children programs that subsidize FV purchases in SNAP households (Verghese et al. 2019).

The provision of free FV to those seeking food assistance has been included in past White House budget proposals (Harvest Box proposal, see Karst 2020) and in USDA’s plan to bolster food assistance in response to COVID-19 (USDA 2020a). Free and subsidized FV are also elements of emerging ‘Food as Medicine’ platforms (Mozaffarian et al. 2022), which include medically tailored grocery and produce prescription programs in which health care professionals leverage healthcare funds to provide access to or delivery of foods such as FV that support treatment of or prevention of diet-related health conditions (e.g., diabetes). This includes the US Centers for Medicare and Medicaid Services (CMS), which has permitted states to experiment with coverage of certain food-based interventions since 2010 (Hanson et al. 2024), and several private health systems that have tested produce prescriptions with calls for increased future federal funding (Adams et al. 2024).

Simultaneously the federal government pursues ambitious plans to cut domestic food waste. The Obama administration set a goal to cut waste in half by 2030, and the Trump (US EPA, 2018) and Biden (USDA 2024) administrations affirmed this strategic direction. Such diverse administrations have aligned to focus on food waste reduction because achieving this goal can enhance food security, reduce environmental and resource burdens, and improve financial outcomes across the food system to the benefit of a broad array of stakeholders. The U.S. national strategy (USDA 2024) explicitly emphasizes actions that “…share food waste prevention tactics— such as food storage or meal planning…” Given that FV are the most wasted category of food and constitute 30-40% of edible food that is wasted by U.S. households (Hoover and Moreno 2017, Li et al. 2023), this implicitly urges households to reconsider FV purchases, preparation, eating patterns and storage to minimize waste.

Given that FV are the most wasted food category (Hoover and Moreno 2017, Li et al. 2023), and given that FV subsidies have been shown to have mixed impacts on FV expenditures and intake (Smith-Drelich 2016), there have been calls for research on the interaction between the subsidization, intake, and waste of FV in order to ensure FV consumption incentive programs are designed to minimize FV waste (Burgaz et al. 2023, Xie et al. 2021, Burrington et al. 2020, Metcalfe et al. 2022) and to ensure simulations that explore the simultaneous impact of such policies on food security and food waste are appropriately calibrated (Mason-D’Croz et al. 2019).

The purpose of this article is to report the findings of a randomized trial in which participants pick up free FV sufficient to meet ∼60% of household needs from a centralized location for three consecutive weeks. Half were randomized to a smart tailored coaching intervention that emphasizes reducing food waste and the replacement of less nutritious foods with FV (hereafter, treatment) while the remaining participants were randomized to an intensity-matched tailored coaching intervention focused on stress management (hereafter, control). Participating households measured their FV inventory before and after the study while detailed food acquisition, intake and waste data is collected via the FoodImage app (Roe et al. 2020) for three consecutive days before and after all free FV pickups occur.

## Methods

### Intervention and Study design

The study is a randomized trial conducted over a 4-week period as detailed in Figure 1. The study design was registered at ClinicalTrials.gov (NCT05061888) prior to study commencement. The current study was approved by the Institutional Review Board of the Pennington Biomedical Research Center (Protocol: 2021-015-PBRC), and all participants provided written informed consent.

**Figure 1.**
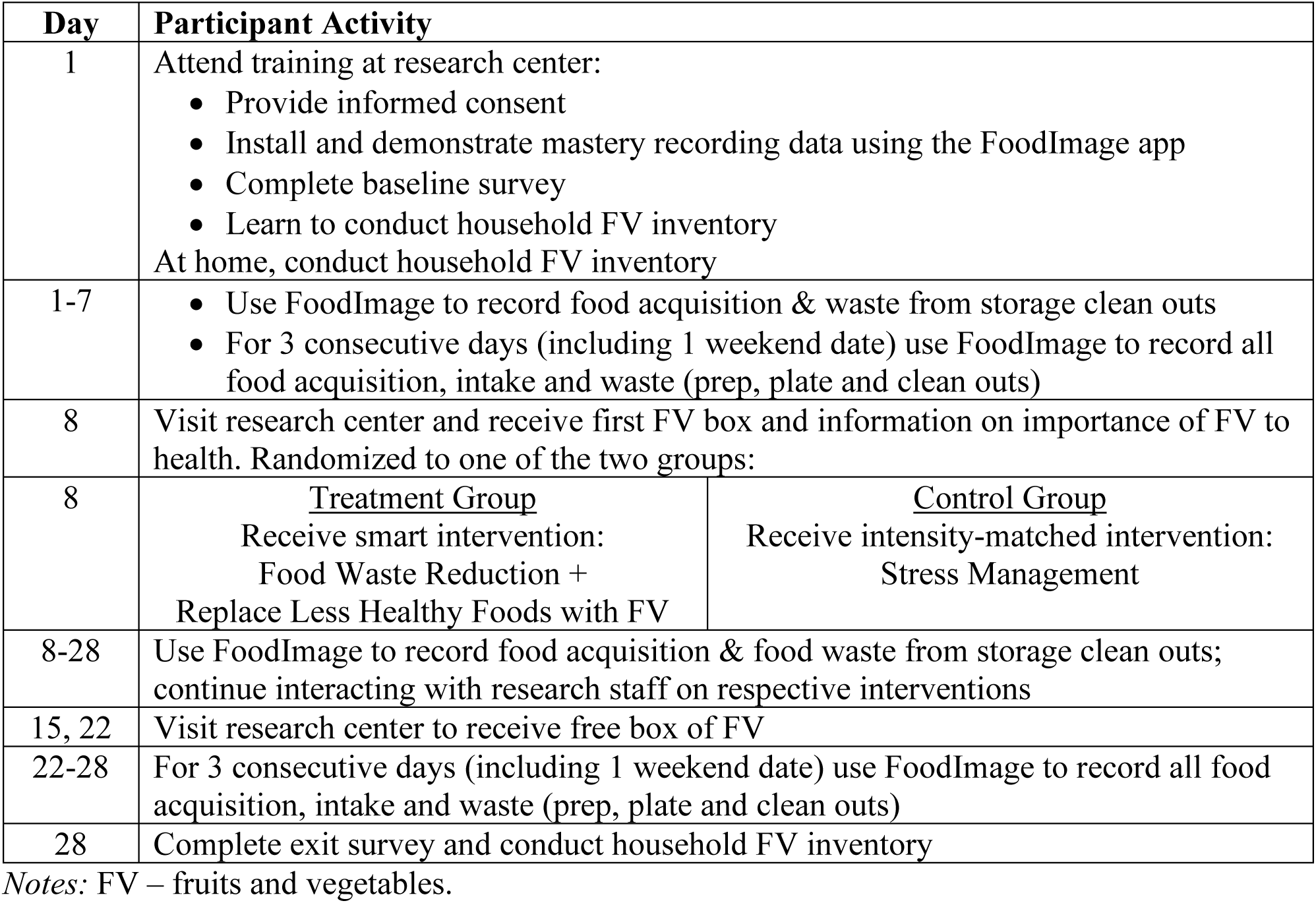
Study Activities.

Recruited participants attended an individual session where, after completing a baseline survey in REDCap, staff verified proper installation of the FoodImage app and provided training until participants demonstrated mastery in using the app to record and transmit data. Participants learned to conduct a home inventory of FV, which they completed after returning home with results transmitted to research staff that same day and then repeated at the end of the study.

All participants recorded each instance of food acquisition (not reported herein) and food discards from storage clean outs (‘toss’ events) for the duration of the study. In addition, during the first seven days all participants recorded data about discards that occurred during food preparation events (‘prep’ events) and meals (‘eat’ events) for at least 3 continuous days (including at least one weekend day). As appropriate to the focal event, recorded data included photos of food shopping receipts (shop), food preparation waste (prep), food consumption (photo of plates prior to and upon completion of each eating occasion both at home and while dining out with intake being the difference), and purges of uneaten food from storage (toss). For participants in multi-person households, data concerning food shopping, preparation and storage purges may capture household-wide activity, while eating occasion data (food selection, consumption and plate waste) were captured by the participant for each household member who joined the study participant for a given meal. This means that all daily meals (selection, intake and plate waste) of the direct study participant are captured while only some of the daily meals of other household members are captured as other household members may not have joined every daily eating occasion for the direct study participant.

Within the app, the participant also entered information about the meal occasion (e.g., breakfast, snack) and the disposition of uneaten food and food scraps (saved for later, thrown in trash), which was appended to the photo and securely sent to secure servers automatically as participants ended interaction with the app. This permitted staff to monitor if participants regularly sent data and to follow up if participants stopped data collection.

After 7 days, and independent of behavior during baseline monitoring, half of the participants were randomized to the treatment intervention with the remainder randomized to the control intervention. All participants returned to the research center on day 8 to receive the first of three free fresh seasonal FV boxes with the other two picked up on days 15 and 22. The box contained about 40% fruit and 60% vegetables and was sufficient to meet 60±5% of the household’s FV needs as recommended by USDA’s MyPlate given to the size and age profile of household members.

On day 8 the treatment group met individually with a trained coach who presented a structured set of materials that included an introduction to food waste, ways that food waste could be reduced over time, and how FV could be substituted for less healthy foods. The coach and participant then engaged in a lifestyle interview in which the coach asked multiple open-ended questions to gather information about the participant’s typical eating, shopping, preparation, storage, and meal habits, including leftover and disposal tendencies. Questions covered the number of people in the household, number of shopping trips, meal planning (or lack thereof), etc.

After basic information on food waste management and approaches to substitute FV for other foods was introduced and the coach acquired an in depth understanding of current habits, the intervention was tailored to target actions expected to yield high incremental impact for current food waste and FV eating habits. The coach and participant reviewed instructional information and then used specific goal setting techniques to make a plan that reduces food waste in the immediate future (i.e., daily in the next week). The coach and participant also collaborated to find a technique that the participant found logistically and financially feasible and efficacious (high self-efficacy).

Specific goal setting techniques such as SMART (Specific, Measurable, Attainable, Reward, Timebound) goals (Doran 1981) were tailored to ensure that the participant had a concrete idea of what to do to enact the plan. The participant then received regular semi-structured tips via text, email or phone (three to four per week with transmission mode based on participant preference) and follow-up interactions with the coach who assessed participant comprehension of the materials and the efficacy of the chosen strategy. Strategies were modified or added based on gathered data and delivered to assist the participant in following through with plans to reduce food waste with the coach providing feedback and encouragement to promote self-efficacy and motivate change. The control group received a similarly tailored intensity-matched (number of topics, contacts and materials) intervention focused on stress management, i.e., devoid of content on food waste reduction or substitution of FV for less nutritious foods.

After the third and final pick up of the free FV box on day 22, all participants again recorded eat and toss event data for up to 3 continuous days (including at least one weekend day). On day 28 all participants completed another home FV inventory and completed an exit survey.

### Participants

Participants were recruited from the Baton Rouge, Louisiana region via e-mail lists, social media posts, and ads on the research center’s recruitment webpage. No data is available concerning the number of eligible individuals that chose not to participate as recruitment materials included open invitations. Inclusion criteria included: 1) ages of 18-62 years, 2) body mass index of 18.5 – 50 kg/m^2^, 3) performed the majority of household food shopping and preparation, and, 4) for households featuring children, children are ages 6-18 years. Exclusion criteria include: 1) not able to use an iPhone, 2) refusal or unable to use the FoodImage smartphone app to collect data in free-living conditions, 3) households that purchase groceries less than 1 time per week, and 4) more than 2 children living in the household.

Participants used their own iPhone during the study (acknowledging that data will be used during the project). An iPhone is necessary as the FoodImage app was programmed only for Apple devices. Exclusion criteria 3 ensures that at least one shopping episode will take place during the baseline study period while exclusion criteria 4 helps ensure that the randomized groups have similar amounts of food waste at baseline since food waste increases with the number of children in a household (Schanes et al. 2018).

Participants were compensated $100 for the successful completion of week 1 and $165 for successful completion of the remaining 21 days (total compensation = $265). Participants also received a free fresh seasonal FV box once per week during the final 21 days.

### Data Preparation

Records collected via the FoodImage app were prepared for analysis using approaches described elsewhere (Roe et al. 2020). Photos collected via the app were viewed by trained staff who were blinded to the participant’s treatment status and did not interact with participants. Staff identified nutrient matches (Standard Reference 28, USDA (2020b) and the Food and Nutrient Database for Dietary Studies, 2019-2020, USDA (2020c)) and estimated the mass of all food prepared, selected, consumed, returned, and discarded at each stage based upon these images using previously validated methods. Served food that was uneaten but saved for later (leftovers) or directed to other people for consumption purposes (shared) was tracked separately and not marked as discarded. Food consumption is expressed in energy (kilocalorie) units for total food intake and in servings when analyzing FV consumption. Food inventory and waste variables are expressed in mass (grams) units.

The data classifies whether each item’s discard was unavoidable (e.g., bones), possibly avoidable (e.g., apple peels), or avoidable (e.g., meat). Avoidable discards are defined as food waste for the purposes of this analysis. FoodImage’s validity and accuracy for measuring consumer food waste (Roe et al. 2020) and evaluating food waste interventions (Roe et al. 2022) has been previously documented.

Waste was recorded at meal events (eat), during food preparation (prep), and during cleanouts of stored food (toss). As prep and toss events are episodic and need not occur at every meal or even every day, these are pooled across the two study periods (‘pre’ or prior to the first pickup of free FV, and ‘post’ for the remainder of the study). The frequency of reported prep and toss waste events within a period and the average amount discarded per waste event is analyzed. These are reported at the household level. Food intake and plate waste is summarized daily for each tracked household member for all days during which meals were recorded by the participant. In all cases the analyses focused on the waste of FV (rather than other food categories) as the treatment intervention’s focus was on ensuring the free FV was consumed rather than discarded.

FV inventory data was recorded by the participants at the beginning and end of the study period and reported in grams. Inventory figures are available for fruits and vegetables separately and separately by the degree of processing with the amount of fresh and frozen FV analyzed. Freezing acquired FV in excess of immediate needs was promoted by study coaches as a food waste reduction technique. Frozen FV does not distinguish between items purchased as frozen and items that were purchased fresh and then frozen by the participant.

### Statistical Analysis

The impact of study interventions was assessed via a linear random effects regression model:

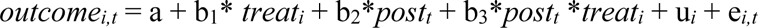

where *outcome_i,t_* is the outcome of interest (e.g., intake, waste, inventory) as recorded for participant *i* at time *t*, *post_t_* = 1 if the data was recorded after the commencement of free FV distribution and = 0 otherwise, *treat_i_* = 1 if the participant was randomly assigned to the treatment group and = 0 otherwise, u*_i_* is the random effects term, which represents the drivers of the outcome variable associated with participant *i* that are unobserved by the research team, and e*_i,t_* represents all other unobserved drivers of the outcome variable associated with participant *i* at time *t*. The parameter b_1_ identifies the effect of free FV pickups on the outcome variable while the parameter b_3_ identifies the differential effect of free FV pickups on the outcome variable for those randomly assigned to the treatment group. Parameter b_2_ identifies any incidental differences in the outcome variable that arises between the control and treatment groups prior to the commencement of free FV pickups. The parameters a, b_1_, b_2_, and b_3_ are estimated using the ‘xtreg’ function in Stata (version 18) where *t*-tests are conducted using robust standard errors to assess whether the parameters are statistically different from zero. The sum of b_2_ and b_3_, which represents the pre/post change in outcome for the treatment group, is also reported and tested. Statistical significance is set at the five percent level with *p-*values between 0.10 and 0.05 deemed marginally significant.

Outliers were defined as an observation that has a studentized residual value greater than 3. Each outlier was individually checked to ensure it was a correctly recorded value and not a reporting error. All outliers identified in the data were verified as correctly reported, and thus no data was removed from any analyses based on its outlier status.

## Results

The 46 participants are balanced between treatment and control on key demographics and initial FV inventories except for initial inventories of frozen FV, which is significantly higher for the control group at baseline (Table 1). The modal sample respondent is in their 40’s, female, white, is employed less than full time, holds a bachelor’s degree, lives in a two-person household, has an annual household income greater than $100,000 and is not currently enrolled for the Supplemental Nutrition Assistance Program (SNAP). The participants feature substantial variation across several characteristics including 30% who identify as Black or African American, 22% who have annual household incomes less than $50,000, 26% who have less than a bachelor’s degree and 17% who are currently receiving SNAP.

**Table 1.**
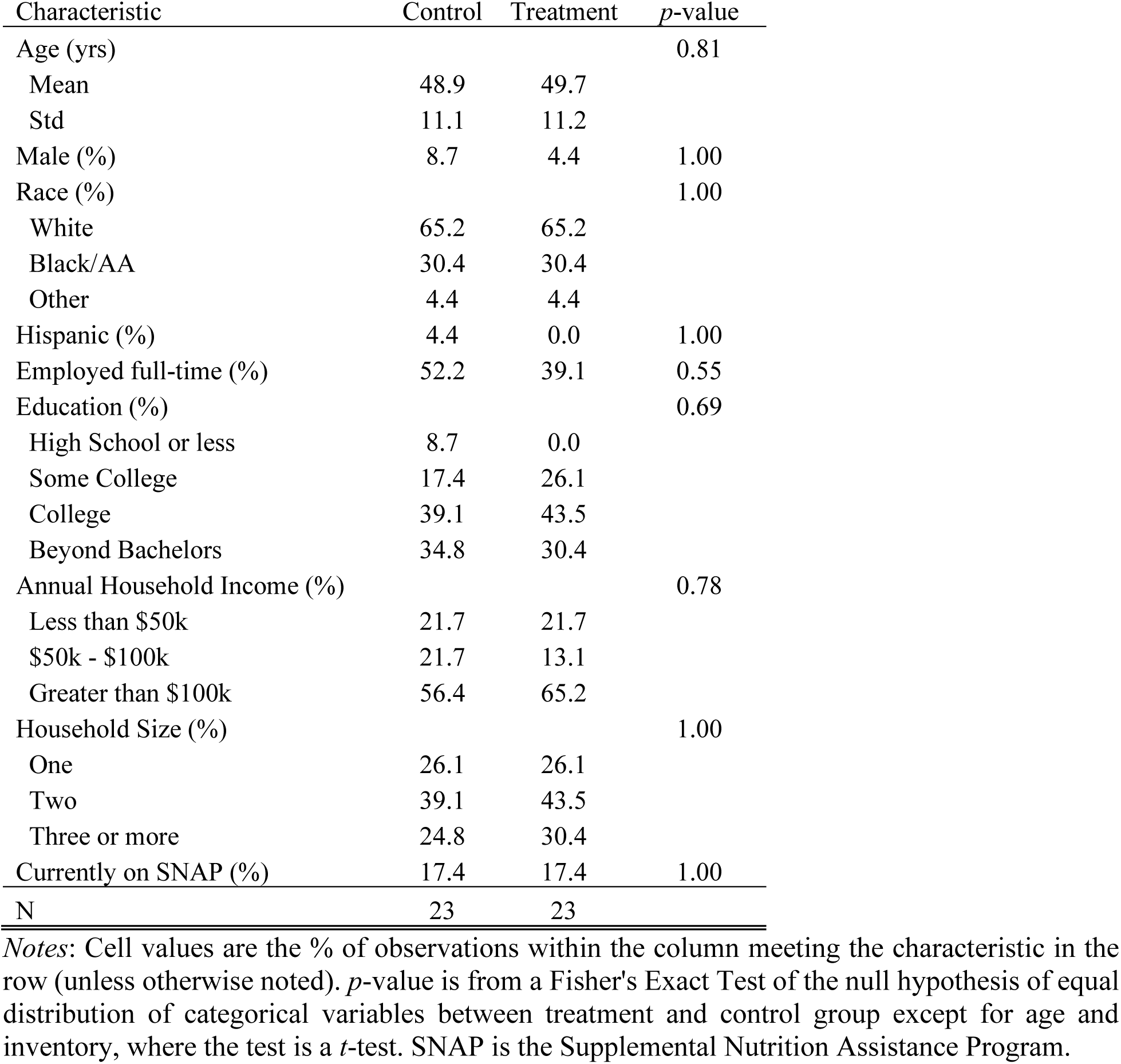
Sample Respondent Characteristics.

| Characteristic | Control | Treatment | *p*-value |
| --- | --- | --- | --- |
| Age (yrs) |  |  | 0.81 |
| Mean | 48.9 | 49.7 |  |
| Std | 11.1 | 11.2 |  |
| Male (%) | 8.7 | 4.4 | 1.00 |
| Race (%) |  |  | 1.00 |
| White | 65.2 | 65.2 |  |
| Black/AA | 30.4 | 30.4 |  |
| Other | 4.4 | 4.4 |  |
| Hispanic (%) | 4.4 | 0.0 | 1.00 |
| Employed full-time (%) | 52.2 | 39.1 | 0.55 |
| Education (%) |  |  | 0.69 |
| High School or less | 8.7 | 0.0 |  |
| Some College | 17.4 | 26.1 |  |
| College | 39.1 | 43.5 |  |
| Beyond Bachelors | 34.8 | 30.4 |  |
| Annual Household Income (%) |  |  | 0.78 |
| Less than \$50k | 21.7 | 21.7 | |
| \$50k - \$100k | 21.7 | 13.1 | |
| Greater than \$100k | 56.4 | 65.2 | |
| Household Size (%) |  |  | 1.00 |
| One | 26.1 | 26.1 |  |
| Two | 39.1 | 43.5 |  |
| Three or more | 24.8 | 30.4 |  |
| Currently on SNAP (%) | 17.4 | 17.4 | 1.00 |
| N | 23 | 23 |  |

*Notes:* Cell values are the % of observations within the column meeting the characteristic in the row (unless otherwise noted). *p*-value is from a Fisher's Exact Test of the null hypothesis of equal distribution of categorical variables between treatment and control group except for age and inventory, where the test is a *t*-test. SNAP is the Supplemental Nutrition Assistance Program.

The mean and standard deviation of the outcome variables are listed in Table 2. Values are provided for both the overall sample and by period and treatment group with differences between groups and periods also listed. In terms of food intake, the study is capturing a study-long average of 1308 kcals across all tracked household members which includes 0.79 servings of fruit and 1.50 servings of vegetables. Plate waste averages 29.3 grams per day while there are 9.5 prep and toss events on average per reporting period that generate 117.4 grams of avoidable food waste per event. FV inventory averaged 31.3 kg per household including 7.5 kg of fresh and 5.8 kg of frozen FV with the remainder consisting of other forms of FV processing (e.g., canned).

**Table 2.** Outcome Variables by Period and Treatment Group.

| **Outcome** | **Pre** | **Post** | **All** | **Difference** |
| --- | --- | --- | --- | --- |
| **Fruit Intake (serv/day)** |  |  |  |  |
| Control | 0.65 (1.04) | 0.72 (1.71) | 0.69 (1.41) | 0.07 |
| Treatment | 0.71 (0.94) | 1.09 (1.28) | 0.90 (1.14) | 0.38 |
| All | 0.67 (0.99) | 0.91 (1.52) | 0.79 (1.29) | 0.24 |
| Difference | 0.06 | 0.37 | 0.21 | 0.31 |
| **Veg Intake (serv/day)** |  |  |  |  |
| Control | 1.48 (1.45) | 1.31 (1.20) | 1.39 (1.33) | -0.17 |
| Treatment | 1.45 (1.09) | 1.78 (1.39) | 1.62 (1.26) | 0.33 |
| All | 1.46 (1.29) | 1.54 (1.32) | 1.50 (1.30) | 0.08 |
| Difference | -0.03 | 0.47 | 0.23 | 0.50 |
| **All food intake (kcal/day)** |  |  |  |  |
| Control | 1390.8 (693.5) | 1246.8 (662.8) | 1318.0 (680.7) | -144.0 |
| Treatment | 1298.6 (691.6) | 1298.9 (722.3) | 1298.7 (706.2) | 0.3 |
| All | 1345.4 (692.8) | 1272.8 (692.4) | 1308.5 (692.9) | -72.6 |
| Difference | -92.2 | 52.1 | -19.3 | 144.3 |
| **All FV Inventory (kg/hh)** |  |  |  |  |
| Control | 30.5 (20.1) | 35.5 (25.7) | 33.0 (22.9) | 5.0 |
| Treatment | 28.1 (26.7) | 31.1 (26.1) | 29.6 (26.2) | 3.0 |
| All | 29.3 (23.4) | 33.3 (25.7) | 31.3 (24.5) | 4.0 |
| Difference | -2.4 | -4.4 | -3.4 | -2.0 |
| **Fresh FV Inventory (kg/hh)** |  |  |  |  |
| Control | 6.2 (5.2) | 10.5 (7.6) | 8.3 (6.8) | 4.3 |
| Treatment | 5.4 (3.3) | 7.8 (4.4) | 6.6 (3.9) | 2.4 |
| All | 5.8 (4.3) | 9.1 (6.2) | 7.5 (5.6) | 3.3 |
| Difference | -0.8 | -2.7 | -1.7 | -1.9 |
| **Frozen FV Inventory (kg/hh)** |  |  |  |  |
| Control | 6.8 (5.4) | 6.6 (6.6) | 6.7 (5.9) | -0.2 |
| Treatment | 4.1 (2.7) | 5.7 (4.2) | 4.9 (3.6) | 1.6 |
| All | 5.4 (4.4) | 6.1 (5.5) | 5.8 (5.0) | 0.7 |
| Difference | -2.7 | -0.9 | -1.8 | 1.8 |
| **FV Plate Waste (g/day)** |  |  |  |  |
| Control | 16.1 (34.5) | 27.8 (70.8) | 22.1 (56.2) | 11.7 |
| Treatment | 41.8 (117.1) | 31.3 (88.4) | 36.4 (103.4) | -10.5 |
| All | 29.0 (87.3) | 29.5 (80.0) | 29.3 (83.6) | 0.5 |
| Difference | 25.7 | 3.5 | 14.4 | -22.2 |
| **Other FV Waste (g/event)** |  |  |  |  |
| Control | 47.4 (90.1) | 163.8 (132.9) | 111.5 (128.6) | 116.4 |
| Treatment | 131.0 (410.8) | 115.4 (146.0) | 122.7 (296.2) | -15.6 |
| All | 91.9 (305.8) | 138.7 (140.7) | 117.4 (231.1) | 46.8 |
| Difference | 83.6 | -48.4 | 11.2 | -132.0 |
| **Other FV Waste (# events)** |  |  |  |  |
| Control | 5.5 (8.1) | 14.3 (12.3) | 10.3 (11.4) | 8.8 |
| Treatment | 5.7 (4.9) | 11.5 (10.9) | 8.8 (9.0) | 5.8 |
| All | 5.6 (6.5) | 12.9 (11.6) | 9.5 (10.2) | 7.3 |
| Difference | 0.2 | -2.8 | -1.5 | -3.0 |

*Notes:* FV plate waste was recorded during ‘eat’ events while other FV waste was recorded during prep and toss events.

The effect of the treatment coaching intervention can be observed in the regression coefficients reported in Table 3. In terms of food intake, the treatment intervention yielded marginally statistically significant increases in fruit (0.33 servings per day, *p* = 0.09) and a statistically significant increase vegetable intake (0.50 servings per day, *p* = 0.010), but no statistically significant change in total caloric intake (*p* = 0.130). We note that the effect of the provision of free FV, independent of whether the respondent received the treatment or control intervention, did reduce total caloric intake (-132.7 kcal/day, *p* = 0.040).

**Table 3.** Regression Model Results of Intervention Impacts.

| **Outcome** | **Initial**<br>**Balance (b<sub>1</sub>)** | **FV Pickup**<br>**Effect (b<sub>2</sub>)** | **FV Pickup x**<br>**Coaching**<br>**Treatment**<br>**(b<sub>3</sub>)** | **b<sub>2</sub> + b<sub>3</sub>** | **# obs**<br>**(R<sup>2</sup>)** |
| --- | --- | --- | --- | --- | --- |
| Fruit Intake<br>(serving/day/p) | -0.00<br>(0.16)<br>[0.99] | 0.09<br>(0.15)<br>[0.55] | 0.33*<br>(0.20)<br>[0.09] | 0.42***<br>(0.13)<br>[0.002] | 530<br>(0.02) |
| Veg Intake<br>(serving/day/p) | -0.06<br>(0.19)<br>[0.74] | -0.17<br>(0.14)<br>[0.24] | 0.50**<br>(0.20)<br>[0.01] | 0.34**<br>(0.14)<br>[0.01] | 530<br>(0.02) |
| All food Intake<br>(kcal/day/p) | -119.3<br>(116.3)<br>[0.30] | -132.7**<br>(64.6)<br>[0.04] | 147.8<br>(97.7)<br>[0.13] | 15.0<br>(73.3)<br>[0.84] | 530<br>(0.005) |
| All FV Inventory<br>(kg/hh) | -2.49<br>(7.01)<br>[0.72] | 4.89**<br>(2.34)<br>[0.04] | -1.83<br>(2.80)<br>[0.51] | 3.06**<br>(1.55)<br>[0.05] | 92<br>(0.01) |
| Fresh FV<br>Inventory<br>(kg/hh) | -0.74<br>(1.28)<br>[0.56] | 4.31***<br>(1.34)<br>[0.001] | -1.96<br>(1.67)<br>[0.24] | 2.36**<br>(1.00)<br>[0.02] | 92<br>(0.12) |
| Frozen FV<br>Inventory<br>(kg/hh) | -2.66**<br>(1.26)<br>[0.04] | 0.16<br>(0.64)<br>[0.80] | 1.78**<br>(0.85)<br>[0.04] | 1.62***<br>(0.56)<br>[0.004] | 92<br>(0.05) |
| FV Plate Waste<br>(g/day/p) | 25.6**<br>(11.3)<br>[0.02] | 11.7*<br>(6.9)<br>[0.09] | -22.2*<br>(13.2)<br>[0.09] | -10.5<br>(11.3)<br>[0.35] | 512<br>(0.01) |
| Other FV Waste<br>(g/event) | 73.4<br>(73.8)<br>[0.32] | 100.5***<br>(36.2)<br>[0.005] | -66.4<br>(41.4)<br>[0.11] | 34.2*<br>(20.0)<br>[0.09] | 103<br>(0.02) |
| Other FV Waste<br>(# events/period) | 0.73<br>(1.92)<br>[0.70] | 9.53***<br>(2.13)<br>[<0.001] | -3.57<br>(3.05)<br>[0.24] | 5.96***<br>(2.19)<br>[0.006] | 103<br>(0.14) |

*Notes:* Each cell in the first three numeric columns lists the parameter estimate from the linear random effects regression model detailed in the text with robust standard error in parentheses and the associated *p*-value in square brackets. The fourth numeric column lists the sum of parameters *b*<sub>2</sub> and *b*<sub>3</sub> and the standard error and *p*-value associated with that linear sum. Parameter estimates that are statistically different from zero at the 10%, 5% and 1% level are denoted by \*, \*\*, \*\*\* respectively. The final column provides the number of observations contributing to the estimated model in a given row and the model's overall R<sup>2</sup> (goodness of fit).

The effect of the treatment intervention on total (*p =* 0.51) and fresh FV inventories (*p* = 0.24) was insignificant, though there was a significant positive impact on frozen FV inventories (1.78 kg/household, *p* = 0.04). The effect of the provision of free FV, independent of intervention received, did increase total (4.89 kg/household, *p* = 0.04) and fresh FV inventories (4.31 kg/household, *p* = 0.001).

In terms of food waste, the intervention led to a marginally significant reduction in FV plate waste (-22.2 g/household/day, *p* = 0.092) but no changes in the number (*p*=0.24) or average magnitude (*p=*0.11) of FV waste events outside of eating occasions (e.g., during preparation or clean out events). The provision of free FV (independent of the intervention) was associated with significantly more FV waste events (9.53 events/period, *p* < 0.001) and greater average FV waste per FV waste event (100.5 g/event, *p* = 0.005), and a marginally significant increase in FV plate waste (11.7 g/person/day, *p* = 0.09).

## Discussion

This study was designed to investigate the dietary and waste effects of providing free FV to households as well as the incremental effect of coupling free FV provision with the treatment coaching intervention focused on replacing less nutritious food options with FV while ensuring the free FV were not wasted. The study respondents’ pre-intervention FV was about 2.1 servings per day, which is below recommendations. In the absence of the treatment intervention, simply providing free FV did not yield significant changes in FV intake among study participants, though total reported caloric intake did decline significantly (by about 10% of baseline reported caloric intake). Among those who received the treatment (rather than the control) intervention, intake of FV did increase by 0.83 servings per day, with a marginally significant increase in fruit (0.33 servings/day) and a statistically significant increase in vegetables (0.50 servings/day). These values exceed the upper bound of 95% confidence intervals documented in a recent meta-analysis (Jabbari et al. 2024), which identifies an increase in aggregate FV intake by 0.55 servings/day (95% CI: 0.34, 0.77), fruit intake by 0.13 (95% CI: -0.01, 0.27), and vegetable intake by 0.15 (95% CI: 0.09, 0.21).

The intervention involved households picking up free FV boxes for three consecutive weeks, which could logically lead to stockpiling. Hence, we measure the impact of free FV provision on the change in household FV inventories. Across both treatment and control participants, there is a statistically significant increase in total (4.89 kg/household, +17%) and fresh FV inventories (4.31 kg/household, +74%) increase over baseline inventories by the end of the study period, but no statistically significant impact on frozen FV inventories. The treatment intervention did yield a statistically significant increase in frozen FV inventories (compared to control) of 1.78 kg/household, representing a 43% average increase over baseline in the frozen FV inventory for treatment households. The treatment intervention had no significant impact on total or fresh FV inventories when compared to the control intervention. The exclusive impact of this intervention on frozen FV inventories aligns with advice given to treatment households that encouraged the freezing of FV received in excess of immediate demand.

The treatment intervention provided advice both on how to increase FV consumption and how to avoid FV waste. However, among both treatment and control participants, the study’s free provision of FV yielded a statistically significantly increase in the number of FV waste events that involved food preparation and food storage cleanout (9.53 events per reporting period) and in the average discard during each of these events (100.5 g/event), though the increase in FV plate waste was only marginally statistically significant (11.7 g/person/day).

The treatment intervention yielded its expected effect with respect to plate waste, with a statistically significant reduction of 22.2 grams per eating occasion versus control participants (a 79% reduction from baseline), but had no significant effects on the number or magnitude of preparation or non-meal discard events compared to the stress management intervention. This mirrors the qualitative pattern of results observed in a randomized control trial reported by Roe et al. (2022), in which the treatment focused solely on food waste reduction and yielded statistically significant reductions in plate waste from all food sources (79%) but no significant changes in preparation or clean out waste events.

As in Roe et al. (2022), it may be that it is more difficult for participants to integrate coaching advice into less frequently occurring events. That it, not every meal involves preparation that lends itself to reducing waste, while shopping and storing activities that can precipitate storage clean outs that generate food waste may be less frequent and more variable, making application of coaching interventions more difficult. The frequency and repetitiveness of many meal occasions may also help explain the relative success of this study’s coaching advice designed to increase FV intake as most meals may have non FV elements that can be replaced with FVs.

Other elements of the study might frustrate attempts to reduce FV waste including that the FV were provided to participants rather than selected by participants. While participant preferences were considered when FV boxes were filled, preferred FVs were not always available, which could exacerbate waste due to some mismatch between preferences and delivered items. Furthermore, the coaching intervention was only 3 weeks, which may limit the ability to change engrained participant habits, particularly as the coaching focus was already split between two outcomes (increasing FV intake and reducing waste).

The study also faces several additional limitations. The sample size is limited in number and in geographic representativeness and is largely female, which impinges upon generalizability. Second, the length of the study was limited. This impedes identifying if excess FV that were added to participants’ frozen FV inventories were eventually consumed or discarded and if short-run FV intake increases were sustained after the coaching intervention stopped. Furthermore, only one form of FV subsidy was considered (free provision requiring pickup at a central location). Future work should explore other forms (e.g., coupons for use at preferred stores). This study also required participants to report to a fixed location to receive free FV; the requirement for a separate trip to obtain the free FV may have altered those who were willing to participate. While nearly 20% of the participants received SNAP during the study, the modal household reported income exceeding $100,000 per year who are unlikely to be targets of governmental or non-profit FV subsidy programs.

## Conclusions

The free or subsidized provision of FV is an increasingly discussed policy for addressing nutritional shortfalls in targeted populations. As these policies require considerable resources, it is imperative to understand how effectively such programs translate resources into increased FV intake and whether FV waste increases, where FV waste represents not only a direct loss of program efficacy as money is spent on items that end up discarded, but also an important indirect societal burden due to the environmental footprint of food that is discarded. We find that the simple provision of free FV representing a substantial portion of a household’s daily FV needs without additional information and support on how to ensure the FV are incorporated into daily meals yields no change in FV intake while increasing the number and magnitude of FV discard events that occur during food preparation and storage cleanout.

The provision of free FV without additional support does yield a statistically significant increase in the household’s ending total and fresh FV inventories. However, these are foods that may be wasted due to their perishable nature and lack of longer-term storage plans. On the other hand, among households who also received intensive coaching about how to substitute less nutritious meal elements with FV and how to avoid food waste, FV intake increased by 0.83 serving per person per day while plate waste declined and ending frozen FV inventories, which may be deployed for later intake, increased. The results of this study reinforce the need to pair interventions along with material support as part of policies and interventions seeking to enhance the intake of nutritious foods while mitigating increases in organic waste.

## Data Availability

All data produced in the present study are available within two years after publication of a peer-reviewed journal article and upon reasonable request to the authors.

## Acknowledgements

The authors acknowledge research support provided by Sarah Ganbat, Mary C. Frances, Isabelle Schexnayder, and Brooklyne Smith. Study data were collected and managed using REDCap electronic data capture tools hosted at Pennington Biomedical Research Center. REDCap (Research Electronic Data Capture, Harris et al. 2009) is a secure, web-based application designed to support data capture for research studies, providing: 1) an intuitive interface for validated data entry; 2) audit trails for tracking data manipulation and export procedures; 3) automated export procedures for seamless data downloads to common statistical packages; and 4) procedures for importing data from external sources.

## References

An, R. (2013). Effectiveness of subsidies in promoting healthy food purchases and consumption: a review of field experiments. Public Health Nutrition, 16(7), 1215–1228.

Adams, E.L., Rigueroa, R., White, K.E., Bell, B.M., Alegria, K., Yaroch, A.L. (2024). Prioritize “Food is Medicine” initiatives in the 2024 Farm Bill for human and planetary health. Translational Behavioral Medicine, 15(6), 330–332. 10.1093/tbm/ibad083

Biener, A. J. Cawley, C. Meyerhoefer. (2017). The high and rising costs of obesity to the US health care system. Journal of General Internal Medicine, 32(Suppl 1):S6–S8. DOI:10.1007/s11606-016-3968-8

Bhat, S., Coyle, D. H., Trieu, K., Neal, B., Mozaffarian, D., Marklund, M., & Wu, J. H. (2021). Healthy food prescription programs and their impact on dietary behavior and cardiometabolic risk factors: a systematic review and meta-analysis. Advances in Nutrition, 12(5), 1944–1956.

Black, A. P., Brimblecombe, J., Eyles, H., Morris, P., Vally, H., & Kerin, O. (2012). Food subsidy programs and the health and nutritional status of disadvantaged families in high income countries: a systematic review. BMC Public Health, 12(1), 1099.

Burgaz, C., Gorasso, V., Achten, W. M., Batis, C., Castronuovo, L., Diouf, A., Asiki, G., Swinburn B.A., Unar-Munguía M., Devleesschauwer B, Sacks G. & Vandevijvere, S. (2023). The effectiveness of food system policies to improve nutrition, nutrition-related inequalities and environmental sustainability: A scoping review. Food Security, 15(5), 1313–1344.

Burrington, C. M., Hohensee, T. E., Tallman, N., & Gadomski, A. M. (2020). A pilot study of an online produce market combined with a fruit and vegetable prescription program for rural families. Preventive Medicine Reports, 17, 101035.

Doran, G. T. (1981). There’s a SMART way to write management’s goals and objectives. Management Review, 70(11), 35–36.

Gittelsohn, J., Trude, A. C. B., & Kim, H. (2017). Peer Reviewed: Pricing Strategies to Encourage Availability, Purchase, and Consumption of Healthy Foods and Beverages: A Systematic Review. Preventing Chronic Disease, 14, E107. DOI: 10.5888/pcd14.170213.

Hanson, E., Albert-Rozenberg, D., Garfield, K. M., Broad Leib, E., Ridberg, R. A., Hager, K., & Mozaffarian, D. (2024). The evolution and scope of Medicaid Section 1115 demonstrations to address nutrition: a US survey. Health Affairs Scholar, 2(2), qxae013.

Harris, P.A., Taylor, R., Thielke, R. Payne, J. Gonzalez, N. Conde, J.G. (2009). Research electronic data capture (REDCap) - A metadata-driven methodology and workflow process for providing translational research informatics support. Journal of Biomedical Informatics 42(2):377–81.

Hoover, D., & Moreno, L. (2017). Estimating quantities and types of food waste at the city level. Natural Resources Defense Council. Report R-17-09-B, October. Available online at: https://www.nrdc.org/sites/default/files/food-waste-city-level-report.pdf (accessed 21 April 2020).

Jabbari M, Namazi N, Irandoost P, Rezazadeh L, Ramezani-Jolfaie N, Babashahi M, Pourmoradian S, Barati M. (2024). Meta-analysis of community-based interventions on fruits and vegetables consumption in adults. Nutrition & Food Science, 54(1),164–91.

Karst, T. (2020). Budget proposal for harvest box draws retail ire. Farm Journal, Feb. 12. Available online at: https://www.agweb.com/article/budget-proposal-harvest-box-draws-retail-ire

Lee, S. H., Moore, L. V., Park, S., Harris, D. M., & Blanck, H. M. (2022). Adults meeting fruit and vegetable intake recommendations—United States, 2019. Morbidity and Mortality Weekly Report, 71(1), 1.

Li, R., Shu, Y., Bender, K. E., & Roe, B. E. (2023). Household food waste trending upwards in the United States: Insights from a National Tracking Survey. Journal of the Agricultural and Applied Economics Association, 2(2), 306–317.

Lowery, C. M., Henderson, R., Curran, N., Hoeffler, S., De Marco, M., & Ng, S. W. (2022). Grocery purchase changes were associated with a North Carolina COVID-19 food assistance incentive program: Study examines a COVID-19 food assistance incentive program in North Carolina. Health Affairs, 41(11), 1616–1625.

Mason-D’Croz, D., Bogard, J. R., Sulser, T. B., Cenacchi, N., Dunston, S., Herrero, M., & Wiebe, K. (2019). Gaps between fruit and vegetable production, demand, and recommended consumption at global and national levels: an integrated modelling study. The Lancet Planetary Health, 3(7), e318–e329.

Metcalfe, J. J., McCaffrey, J., Schumacher, M., Kownacki, C., & Prescott, M. P. (2022). Community-based nutrition education and hands-on cooking intervention increases farmers’ market use and vegetable servings. Public Health Nutrition, 25(9), 2601–2613.

Miller, D. L., Schwartz, M. B., & Brownell, K. D. (2019). Primer on US Food and Nutrition Policy and Public Health: Food Sustainability. American Journal of Public Health, 09, 986–988, 10.2105/AJPH.2019.305071.

Mozaffarian, D., Blanck, H. M., Garfield, K. M., Wassung, A., & Petersen, R. (2022). A Food is Medicine approach to achieve nutrition security and improve health. Nature Medicine, 28(11), 2238–2240.

Pollard, J., Kirk, S. L., & Cade, J. E. (2002). Factors affecting food choice in relation to fruit and vegetable intake: a review. Nutrition Research Reviews, 15(2), 373–387.

Roe, B.E., D. Qi, R.A. Beyl, K. E. Neubig, C. K. Martin, J. W. Apolzan. (2020). The validity, time burden, and user satisfaction of the FoodImage^tm^ smartphone app for food waste measurement versus diaries: A randomized crossover trial.” (accepted) Resources, conservation & recycling.

Roe, B. E., Qi, D., Beyl, R. A., Neubig, K. E., Apolzan, J. W., & Martin, C. K. (2022). A randomized controlled trial to address consumer food waste with a technology-aided tailored sustainability intervention. *Resources*, Conservation and Recycling, 179, 106121.

Schanes, K., Dobernig, K., & Gözet, B. (2018). Food waste matters-A systematic review of household food waste practices and their policy implications. Journal of cleaner production, 182, 978–991.

Smith-Drelich, N. (2016). Buying health: assessing the impact of a consumer-side vegetable subsidy on purchasing, consumption and waste. Public Health Nutrition, 19(3), 520–529.

Steer, K. J., Olstad, D. L., Campbell, D. J., Beall, R., Se’era, M. A., Caron-Roy, S., & Spackman, E. (2023). The impact of providing material benefits to improve access to food on clinical parameters, dietary intake, and household food insecurity in people with diabetes: a systematic review with narrative synthesis. Advances in Nutrition, 14(5), 1067–1084.

US Department of Agriculture. (2019). The Gus Schumacher Nutrition Incentive Program: 2019 Request for Applications, National Institute of Food and Agriculture, May 9, Washington, DC.

US Department of Agriculture. (2020a). USDA announces coronavirus food assistance program. Press release #0222.20, April 17. Available online at: https://www.usda.gov/media/press-releases/2020/04/17/usda-announces-coronavirus-food-assistance-program(accessed 21 April 2020).

US Department of Agriculture. (2020b). National Nutrient Database for Standard Reference, Release 28. US Department of Agriculture, Agricultural Research Service, Nutrient Data Laboratory. [cited on: 9 February 2012]. Available from: http://www.ars.usda.gov/ba/bhnrc/ndl

U.S. Department of Agriculture, Agricultural Research Service. (2020c). 2019-2020 Food and Nutrient Database for Dietary Studies Documentation. Available online at: https://www.ars.usda.gov/ARSUserFiles/80400530/pdf/fndds/2019_2020_FNDDS_Doc.pdf (accessed 20 April 2025).

U.S. Department of Agriculture (2024). “Biden-Harris Administration Announces National Strategy to Reduce Food Loss and Waste and Recycle Organics,” Press Release, June 12, available online at: https://www.usda.gov/media/press-releases/2024/06/12/biden-harris-administration-announces-national-strategy-reduce-food (accessed 28 June 2024).

U.S. Department of Agriculture and U.S. Department of Health and Human Services (2020). Dietary Guidelines for Americans, 2020–2025. 9th ed. Washington, DC: US Government Publishing Office. Available online at: DietaryGuidelines.gov (accessed 28 June 2024).

U.S. Environmental Protection Agency. (2018). Trump Administration Launches “Winning on Reducing Food Waste” Initiative. Press release, Oct. 18. Available online at: https://www.epa.gov/newsreleases/trump-administration-launches-winning-reducing-food-waste-initiative (accessed 21 April 2020).

Verghese, A., Raber, M., & Sharma, S. (2019). Interventions targeting diet quality of Supplemental Nutrition Assistance Program (SNAP) participants: A scoping review. Preventive Medicine, 119, 77–86.

Xie, J., Price, A., Curran, N., & Østbye, T. (2021). The impact of a produce prescription programme on healthy food purchasing and diabetes-related health outcomes. Public Health Nutrition, 24(12), 3945–3955.

Yeh, M. C., Ickes, S. B., Lowenstein, L. M., Shuval, K., Ammerman, A. S., Farris, R., & Katz, D. L. (2008). Understanding barriers and facilitators of fruit and vegetable consumption among a diverse multi-ethnic population in the USA. Health Promotion International, 23(1), 42–51.

